# COVID-19 infection and attributable mortality in UK care homes: Cohort study using active surveillance and electronic records (March-June 2020)

**DOI:** 10.1101/2020.07.14.20152629

**Authors:** Peter F Dutey-Magni, Haydn Williams, Arnoupe Jhass, Greta Rait, Fabiana Lorencatto, Harry Hemingway, Andrew Hayward, Laura Shallcross

## Abstract

**Background:** Epidemiological data on COVID-19 infection in care homes are scarce. We analysed data from a large provider of long-term care for older people to investigate infection and mortality during the first wave of the pandemic.

**Methods:** Cohort study of 179 UK care homes with 9,339 residents and 11,604 staff.We used manager-reported daily tallies to estimate the incidence of suspected and confirmed infection and mortality in staff and residents. Individual-level electronic health records from 8,713 residents were used to model risk factors for confirmed infection, mortality, and estimate attributable mortality.

**Results:** 2,075/9,339 residents developed COVID-19 symptoms (22.2% [95% confidence interval: 21.4%; 23.1%]), while 951 residents (10.2% [9.6%; 10.8%]) and 585 staff (5.0% [4.7%; 5.5%]) had laboratory-confirmed infections. The incidence of confirmed infection was 152.6 [143.1; 162.6] and 62.3 [57.3; 67.5] per 100,000 person-days in residents and staff respectively. 121/179 (67.6%) care homes had at least one COVID-19 infection or COVID-19-related death. Lower staffing ratios and higher occupancy rates were independent risk factors for infection.

217/607 residents with confirmed infection died (case-fatality rate: 35.7% [31.9%; 39.7%]). Mortality in residents with no direct evidence of infection was two-fold higher in care homes with outbreaks versus those without (adjusted HR 2.2 [1.8; 2.6]).

**Conclusions:** Findings suggest many deaths occurred in people who were infected with COVID-19, but not tested. Higher occupancy and lower staffing levels were independently associated with risks of infection. Protecting staff and residents from infection requires regular testing for COVID-19 and fundamental changes to staffing and care home occupancy.

## Background

Globally the number of COVID-19 cases continues to increase, with substantially higher rates of infection reported in care homes.^1^ In the UK, an estimated 400,000 residents live in approximately 11,000 care homes for older people, which provide residential care with or without on-site nursing.^2,3^ Care home residents are particularly vulnerable to COVID-19 due to older age, high prevalence of comorbidity,^4^ and frequent exposure to infection through contact with staff, other residents and contaminated surfaces. At the peak of the pandemic, deaths recorded in UK care homes were three times higher than during the preceding year.^5^ Staff also had higher aged-standardised rates of COVID-19 related mortality compared to other occupations.^6^ UK statistics suggests two-thirds of excess deaths recorded in residents in the last 6 months involved COVID-19,^5^ but this is likely to be an underestimate because many residents were not tested. Understanding the proportion of excess deaths that can be directly and indirectly attributed to COVID-19 infection is important, to fully assess the impact of the pandemic on care homes.

Strategies to protect residents and staff from SARS-CoV-2 include rapid testing, restriction of visitors, and vaccination. These require knowledge of the burden of and risk factors for infection in residents and staff in care homes, linked to outcomes, which may only be drawn from evidence from the pandemic’s first wave. Population-based prevalence surveys and studies based on routine data have demonstrated variation in the incidence of infection and case-fatality between countries,^7–9^ but many people with symptoms were not tested, particularly at the start of the pandemic due to limited testing capacity. There is no syndromic surveillance for infection in care homes in England, and widespread regular testing for SARS-CoV-2 using reverse transcriptase polymerase chain reaction (RT-PCR) was not established for staff and residents until 11 May 2020.^10^ Prior to this, testing was only available for residents or staff who were admitted to hospital, or as part of Public Health England’s outbreak investigations which permitted a maximum of five tests per care home. Consequently, national estimates of incidence and prevalence based on the first wave of infection (February–July 2020) substantially underestimate the burden of infection in care home residents and staff.

To our knowledge, there are no studies which have employed population-level active surveillance (daily monitoring to identify possible cases of COVID-19 in residents and staff) in care homes to investigate the epidemiology and clinical outcomes of both suspected and confirmed COVID-19 infections. We analysed electronic health records from the Four Seasons Health Care Group (FSHCG), one of the UK’s largest for-profit providers of residential and nursing care, with the aim of identifying strategies to protect staff and residents in care homes from future waves of infection. Our objectives were to estimate incidence of and risk factors for infection, and incidence of mortality in the following groups: residents with no evidence of infection; (B) symptomatic residents; (C) asymptomatic residents with confirmed infection; and (D) symptomatic residents with confirmed infection. We also estimated mortality attributable to COVID-19.

## Methods

### Study population and setting

Staff and residents living/working in care homes for older people run by the FSHCG between 2 March and 14 June 2020 were eligible for study inclusion. FSHCG provides a combination of residential and nursing care (for residents with medical conditions), which is predominantly state-funded. Most residents are permanent, but a small proportion receive temporary (respite) care.

In 2020, there were 9,568 beds, representing 9% of all registered care home beds in England, Scotland and Northern Ireland (supplementary methods). 90% of FSHCG homes participated in the whole care home testing programme, implying that all staff and residents were tested for COVID-19 at least once between 11 May and 22 June 2020.

### Data sources

We extracted organisational data, individual-level data for 8,713 residents and aggregate data for all staff and residents (Figure 1). Electronic records collected by the FSHCG are primarily used for billing and monitoring, but have also been used in previous research.^11^

**Figure 1.**
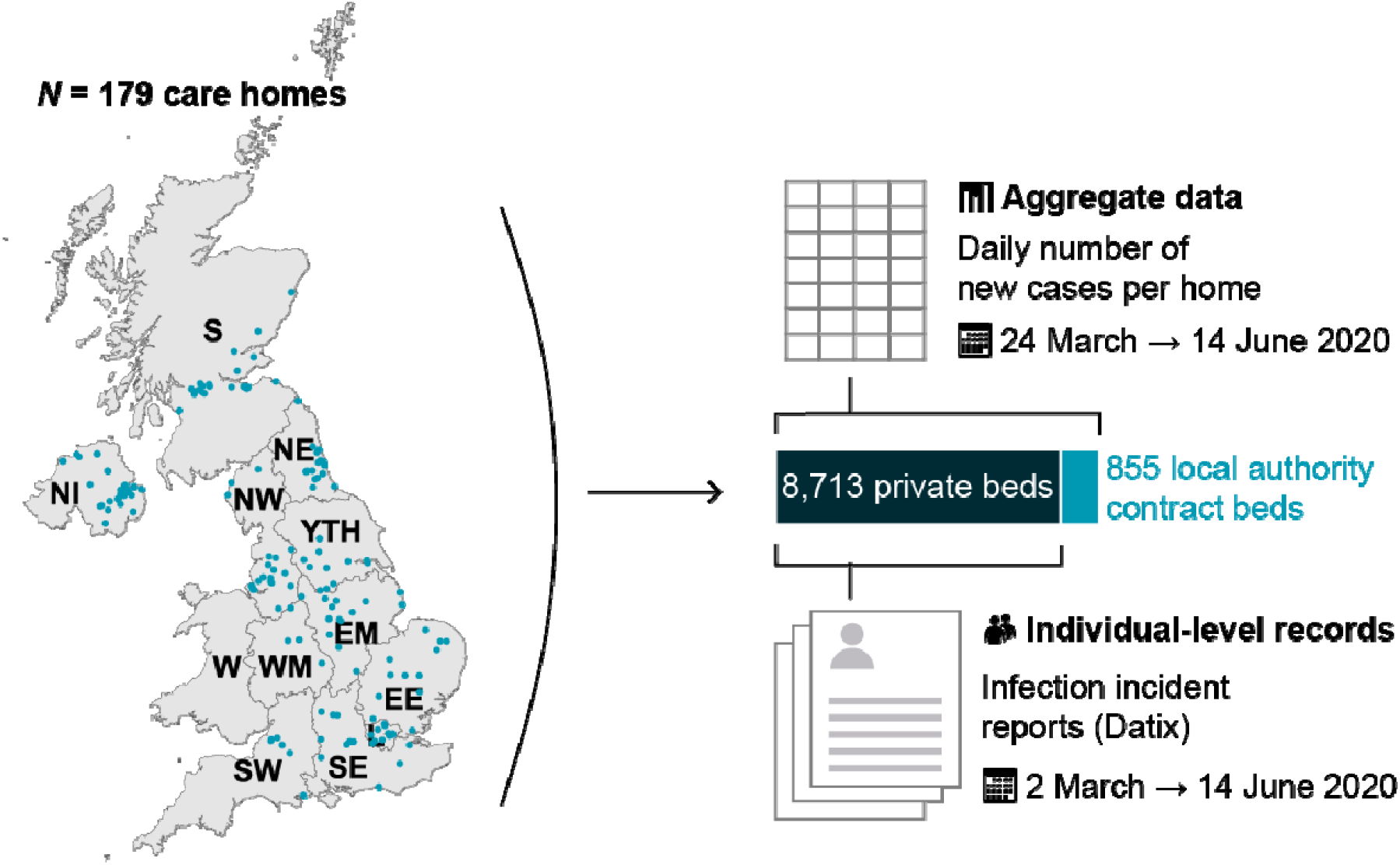
Study overview: location of FSHCG care homes and diagram of data sources *Note:* NI: Northern Ireland; S: Scotland; W: Wales; NE: North East; NW: North West; YTH: Yorkshire and The Humber; EM: East Midlands; WM: West Midlands; EE: East of England; L: London; SE: South East; SW: South West.

#### Individual-level data

FSHCG collects electronic records on residents occupying ‘private’ beds, excluding those occupying beds that are ‘block contracted’ to the local authority (855 beds, see Figure 1). Records include: dates of entry to and exit from the care home, sex, date of birth, type of stay (residential/nursing) and care (general/dementia/older residents). Individual-level data on incidents including infections are reported via ‘Datix’: resident names, care home identifier, incident date/time, date of birth, sex, COVID-19 symptoms (9 multiple choices), test results, resident current location (care home/hospital), and death. Individual-level data on residents were linked to Datix reports (supplementary methods), and used to categorise residents’ infection status into four groups: (A) residents with no evidence of infection (not tested and/or no symptoms); (B) symptomatic residents (symptoms and not tested or tested negative); (C) asymptomatic residents with confirmed infection (no symptoms but tested positive); and (D) symptomatic residents with confirmed infection (symptoms and tested positive) (supplementary methods). The term ‘confirmed’ denoted a positive PCR test. Datix was also used to differentiate deaths in hospital from those in the care home, and to identify COVID-19 related deaths. 1,492/1,880 (79%) of Datix reports were successfully linked.

#### Aggregate data

On 24 March FSHCG introduced a new reporting system requiring managers of each care home to report daily tallies in residents (new symptomatic cases, new confirmed infection in facility, new confirmed infection in hospital, deaths related to COVID-19) and staff (new symptomatic cases, new confirmed cases). The number of occupied beds in each care home was reported weekly via the same mechanism. COVID-19 related deaths were defined as death in a resident with confirmed infection or a death attributed to COVID-19 by the coroner.

### Risk factors

Risk factors included individual-level variables (age, sex, general or dementia care, residential versus nursing care) and care home characteristics (nursing/residential, number of beds, occupancy, bed-to-staff ratio, Index of Multiple Deprivation^12^) obtained from FSHCG. Baseline care home occupancy was computed by averaging weekly occupancy in January-March 2020, before the first COVID-19 case, in order to calculate a ratio of baseline occupancy to the number of bedrooms, and modelled as a continuous variable. We also estimated the ratio of beds to staff as a continuous variable.

A dummy variable marked the time from which an outbreak occurred, defined throughout the manuscript as a care home recording ≥1 confirmed infection or COVID-19 related death. This definition was preferred over a standard outbreak definition (≥2 cases linked in time/place) to compensate for poor COVID-19 case-ascertainment during the pandemic due to limited testing. Sensitivity analysis using a more specific outbreak definition can be found in supplementary methods.

### Statistical analysis

#### Infection in staff and residents in care homes

Incidence and cumulative incidence were calculated for residents and staff using the aggregate daily tallies, the trusted source used for national reporting of cases in all residents and staff (Figure 1).^13^ Daily occupancy and numbers of residents at risk of infection were inferred using interpolation and a life table approach (supplementary methods). The life table allowed us to compute Kaplan-Meier product limit estimators of the cumulative incidence of symptoms, confirmed infections, and COVID-19 related deaths by day based on the aggregate dataset. The incidence rate ratio for care home (based on aggregate data) versus community infections was estimated by contrasting the cumulative incidence for confirmed cases in England with estimates from a national household survey for the period 11 May-7 June 2020.^14,15^

Infection incidence was also estimated from the individual-level dataset, but was subject to under-reporting. Due to this, individual-level data were only used to estimate age/sex-specific rates of infection and Cox proportional hazards models testing the association with individual and organisational-level risk factors.

#### Mortality, attributable mortality and risk factors

The aggregate dataset was used to estimate the crude rate of COVID-19 related mortality in residents. Individual-level data were used to estimate rates of all-cause mortality and case-fatality by age and gender.

To investigate COVID-19 excess mortality, we made the assumption that residents in ‘non-outbreak’ care homes (no record of any confirmed cases or COVID-19 related deaths) had not been exposed to infection, and would therefore not experience excess COVID-19 mortality.^16^ A Cox proportional hazards regression model tested the effect of individual- and home-level risk factors on all-cause mortality, alongside the effect of the time-variant infection status (groups A-D) and care home outbreak status. We estimated the attributable fraction of deaths for each infection category in care homes with and without outbreaks, taking the reference category as individuals with no direct evidence of infection (group A) in non-outbreak care homes. This fraction was obtained by using the model to predict the counterfactual mortality, then computing the attributable fraction within study.^17^ Ninety-five percent confidence intervals for proportions and rates were computed from the exact Poisson and binomial limits. Huber sandwich estimators of variance accounted for the design effect of care home clustering in regression models.

Data were analysed in R3.5.0, epitool^18^ and survival.^19^ Computer scripts are available online.^20^

## Results

### Study population

The study included 9,339 residents across England, Scotland and Northern Ireland and 11,604 staff. 121/179 (67.6%) care homes, totalling 7,102 residents, recorded an outbreak. The mean duration of follow-up for residents and staff was 71 days and 82 days respectively in the aggregate dataset, and 86 days in the individual-level dataset.

### Infection and COVID-19 related mortality (aggregate data)

Care home managers recorded symptoms of infection in 2,075 residents, contributing to an overall cumulative incidence of 22.2% [21.4%; 23.1%] or an incidence rate of 368.0 per 100,000 resident-days [352.3; 384.2] (Table S1a, Figure 2). An additional 951 residents had confirmed infections, of whom 199 were diagnosed in hospital. The cumulative incidence of confirmed infection was 10.2% [9.6%; 10.8%], with an incidence rate of 152.6 per 100,000 [143.1; 162.6]. The rate of confirmed infections in care homes in England was 13-fold higher in care homes compared to the community prevalence of infection derived from the ONS household infection survey (IRR = 12.7 [8.9; 18.3]).^14^

**Figure 2.**
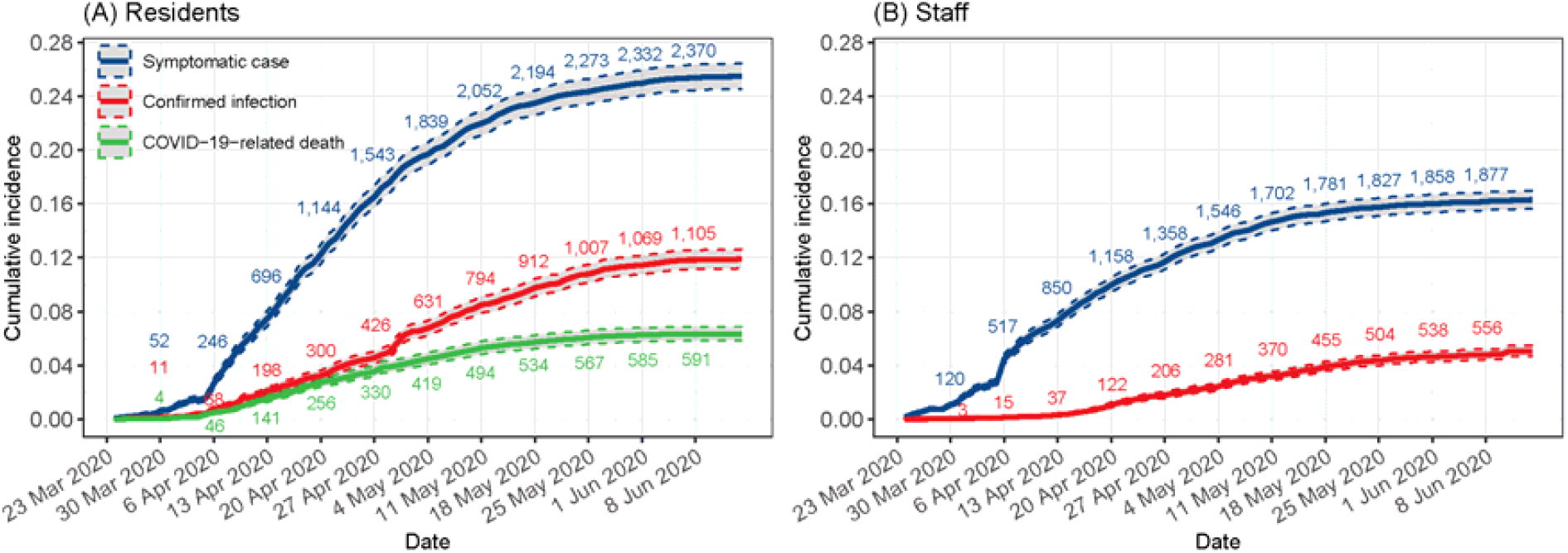
Kaplan-Meier estimates of the cumulative incidence of symptomatic cases, confirmed infections and COVID-related deaths in (A) residents (n=9,339) and staff (n=11,604) according to FSHCG aggregate data (24 Mar 2020-14 Jun 2020) Note: underlying data available on request from authors, subject to permissions from FHSCG.

Care home managers recorded 526 COVID-19 related resident deaths, equivalent to a crude incidence of 5.6% [5.2; 6.1] or 79.7 [73.0; 86.8] per 100,000 resident-days. 24.7% of these deaths took place in hospital (Table S1a).

Care home managers recorded 1,892/11,604 staff (16.3% [15.6%; 17.0%]) experiencing symptoms of infection during the study period, while 585 (5.0% [4.7%; 5.5%]) had a confirmed infection (Table S2, Figure 2).

### All-cause mortality (individual-level data)

Individual-level data were available for 8,713 (93.3%) private residents (Table 3), who accounted for 1,694 all-cause deaths, equivalent to a crude cumulative incidence of 19.4% [18.6%; 20.3%]. The proportion of resident deaths was two-fold higher in care homes with outbreaks compared to those without outbreaks (22.6% versus 11.2%).

**Table 3.** Characteristics of FSHCG private bed residents by type of care home, sex, age, region and status on study exit (2 Mar 2020-14 Jun 2020)

|  | **Outbreak homes**<br>**(N=6328)** | **Other homes**<br>**(N=2385)** | **Total**<br>**(N=8713)** |
| --- | --- | --- | --- |
| **Sex** |  |  |  |
| Female | 4051 (64.0%) | 1616 (67.8%) | 5667 (65.0%) |
| Male | 2277 (36.0%) | 769 (32.2%) | 3046 (35.0%) |
| **Age** |  |  |  |
| <75 years | 1069 (16.9%) | 355 (14.9%) | 1424 (16.3%) |
| 75–84 years | 2113 (33.4%) | 752 (31.5%) | 2865 (32.9%) |
| 85–94 years | 2577 (40.7%) | 1052 (44.1%) | 3629 (41.7%) |
| 95+ years | 569 (9.0%) | 226 (9.5%) | 795 (9.1%) |
| **Resident type** |  |  |  |
| General/elderly | 3799 (60.0%) | 1495 (62.7%) | 5294 (60.8%) |
| Dementia | 2529 (40.0%) | 890 (37.3%) | 3419 (39.2%) |
| **Admission type** |  |  |  |
| Continuing care/independent living | 293 (4.6%) | 58 (2.4%) | 351 (4.0%) |
| Permanent | 5375 (84.9%) | 2065 (86.6%) | 7440 (85.4%) |
| Respite | 660 (10.4%) | 262 (11.0%) | 922 (10.6%) |
| **Funding type** |  |  |  |
| Residential | 1992 (31.5%) | 742 (31.1%) | 2734 (31.4%) |
| Nursing | 4336 (68.5%) | 1643 (68.9%) | 5979 (68.6%) |
| **Infection status by 14 June** |  |  |  |
| Uninfected | 5268 (83.2%) | 2274 (95.3%) | 7542 (86.6%) |
| Symptomatic (not confirmed) | 453 (7.2%) | 111 (4.7%) | 564 (6.5%) |
| Asymptomatic confirmed | 133 (2.1%) | 0 (0.0%) | 133 (1.5%) |
| Symptomatic confirmed | 474 (7.5%) | 0 (0.0%) | 474 (5.4%) |
| **Status as of 14 June** |  |  |  |
| Deceased | 1428 (22.6%) | 266 (11.2%) | 1694 (19.4%) |
| In Home | 4558 (72.0%) | 2011 (84.3%) | 6569 (75.4%) |
| Permanently Discharged | 215 (3.4%) | 69 (2.9%) | 284 (3.3%) |
| Temporary Discharged | 127 (2.0%) | 39 (1.6%) | 166 (1.9%) |
| **Region/nation** |  |  |  |
| East Midlands | 333 (5.3%) | 285 (11.9%) | 618 (7.1%) |
| East of England | 338 (5.3%) | 274 (11.5%) | 612 (7.0%) |
| London | 619 (9.8%) | 0 (0.0%) | 619 (7.1%) |
| North East | 821 (13.0%) | 197 (8.3%) | 1018 (11.7%) |
| North West | 965 (15.2%) | 120 (5.0%) | 1085 (12.5%) |
| Northern Ireland | 1054 (16.7%) | 770 (32.3%) | 1824 (20.9%) |
| Scotland | 785 (12.4%) | 449 (18.8%) | 1234 (14.2%) |
| South East | 567 (9.0%) | 26 (1.1%) | 593 (6.8%) |
| South West | 171 (2.7%) | 71 (3.0%) | 242 (2.8%) |
| West Midlands | 105 (1.7%) | 127 (5.3%) | 232 (2.7%) |
| Yorkshire and The Humber | 570 (9.0%) | 66 (2.8%) | 636 (7.3%) |
| **Index of Multiple Deprivation** |  |  |  |
| 1 - least deprived | 490 (7.7%) | 447 (18.7%) | 937 (10.8%) |
| 2 | 964 (15.2%) | 544 (22.8%) | 1508 (17.3%) |
| 3 | 1946 (30.8%) | 374 (15.7%) | 2320 (26.6%) |
| 4 | 1221 (19.3%) | 459 (19.2%) | 1680 (19.3%) |
| 5 - most deprived | 1707 (27.0%) | 561 (23.5%) | 2268 (26.0%) |

217 deaths occurred in residents with confirmed infection, equivalent to an all-cause case-fatality rate in infected residents (Groups C and D) of 35.7% [31.9%; 39.7%] (Table S4). The case-fatality rate increased with age and was higher in men.

### Factors associated with confirmed infections (individual-level data)

Factors affecting rates of confirmed infections were investigated in Cox Proportional Hazard models (Table 5). Male sex, age ≥85 years, and nursing care (adjusted hazard ratio HR = 1.6 [1.0; 2.4]) were all independently associated with increased risk of confirmed infection. After controlling for organisational differences, care home size no longer had a statistically significant association with rates of infection (adjusted HR = 1.7 [0.7; 4.3] for care homes with ≥70 beds versus <35 beds). Care home baseline occupancy and staffing ratios had the greatest effect on residents’ risk of infection. For example, a 10 percentage point increase in the ratio of occupants to bedrooms was associated with a 51% increase in infection (adjusted HR = 1.5 [1.1; 2.1]); a 10 percentage point increase in the ratio of beds to staff was associated with a 26% increase in infection (adjusted HR = 1.3 [1.1; 1.5]).

**Table 5.**
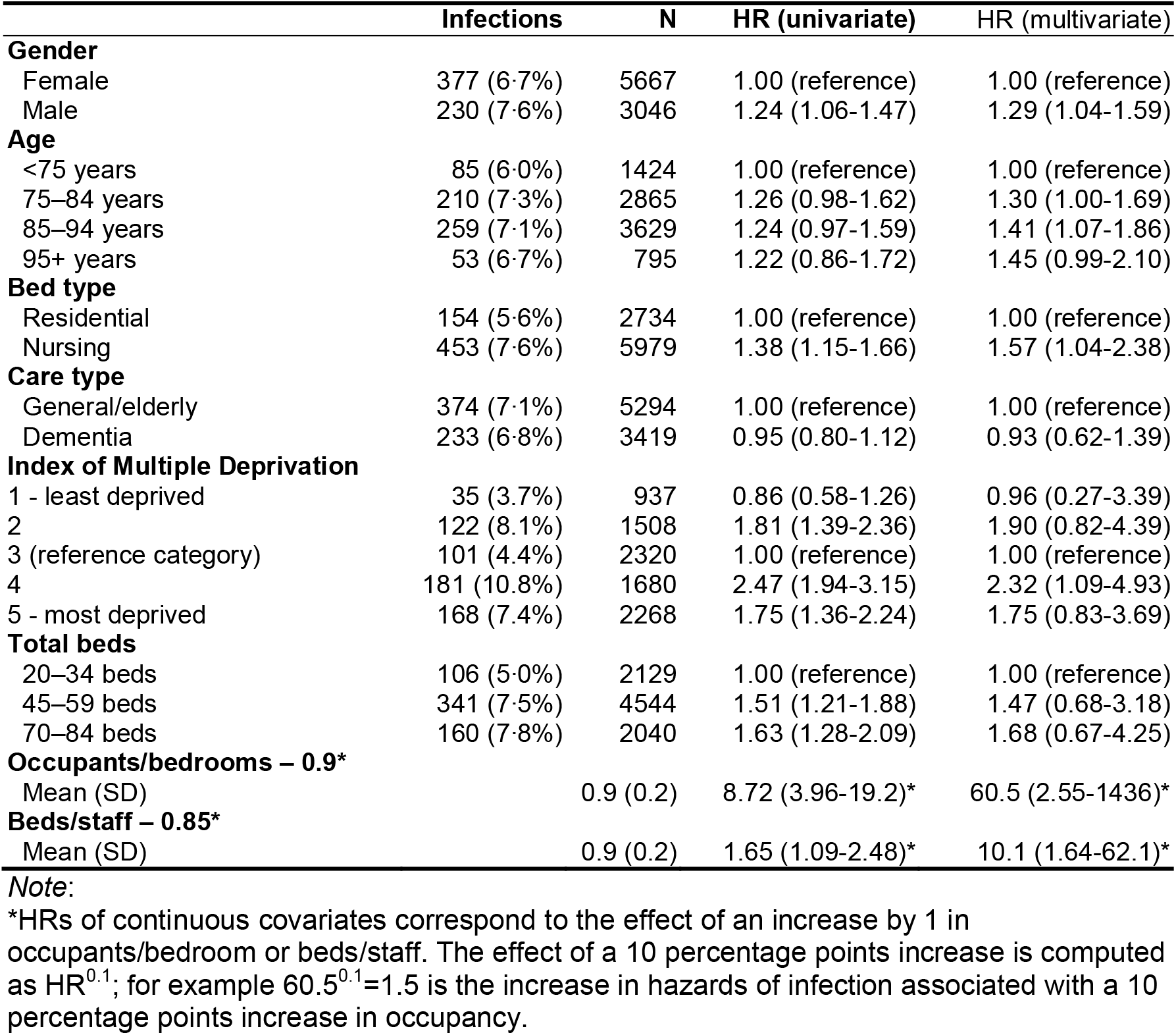
Risk factors for confirmed infection in private residents: hazard ratios (HR) from a Cox proportional hazards model (n=8,713)

### Factors associated with all-cause mortality (individual-level data)

Time-dependent Cox proportional hazard models (Table 6) examined the relationship between infection status (groups A–D) and mortality (Figure S3). After controlling for other risk factors, increased mortality was independently associated with older age, male gender (adjusted HR = 1.5 [1.3; 1.6]), and nursing care (adjusted HR = 1.3 [1.1; 1.6]).

**Table 6.** Risk factors for all-cause mortality in private residents of care homes with and without COVID-19 outbreaks: hazard ratios (HR) from a Cox proportional hazards model (n=8,713, 2 Mar 2020-14 Jun 2020)

|  | Deaths | N | HR (univariate) | HR (multivariate) |
| --- | --- | --- | --- | --- |
| **Gender** |  |  |  |  |
| Female | 996 (17.6%) | 5667 | 1.00 (reference) | 1.00 (reference) |
| Male | 698 (22.9%) | 3046 | 1.40 (1.27-1.54) | 1.46 (1.32-1.61) |
| **Age** |  |  |  |  |
| <75 years | 201 (14.1%) | 1424 | 1.00 (reference) | 1.00 (reference) |
| 75–84 years | 520 (18.2%) | 2865 | 1.30 (1.11-1.53) | 1.35 (1.13-1.62) |
| 85–94 years | 761 (21.0%) | 3629 | 1.50 (1.28-1.75) | 1.73 (1.47-2.03) |
| 95+ years | 212 (26.7%) | 795 | 1.93 (1.59-2.35) | 2.32 (1.86-2.89) |
| **Bed type** |  |  |  |  |
| Residential | 420 (15.4%) | 2734 | 1.00 (reference) | 1.00 (reference) |
| Nursing | 1274 (21.3%) | 5979 | 1.38 (1.24-1.54) | 1.34 (1.12-1.61) |
| **Care type** |  |  |  |  |
| General/elderly | 1015 (19.2%) | 5294 | 1.00 (reference) | 1.00 (reference) |
| Dementia | 679 (19.9%) | 3419 | 1.02 (0.92-1.12) | 1.02 (0.88-1.19) |
| **Index of Multiple Deprivation** |  |  |  |  |
| 1 - least deprived | 469 (50.1%) | 937 | 0.99 (0.84-1.17) | 1.05 (0.79-1.40) |
| 2 | 190 (12.6%) | 1508 | 0.85 (0.73-0.99) | 0.86 (0.65-1.15) |
| 3 (reference category) | 266 (11.5%) | 2320 | 1.00 (reference) | 1.00 (reference) |
| 4 | 321 (19.1%) | 1680 | 0.92 (0.80-1.06) | 0.87 (0.66-1.13) |
| 5 - most deprived | 448 (19.8%) | 2268 | 0.98 (0.86-1.12) | 0.87 (0.68-1.11) |
| **Total beds** |  |  |  |  |
| 20–34 beds | 373 (17.5%) | 2129 | 1.00 (reference) | 1.00 (reference) |
| 45–59 beds | 872 (19.2%) | 4544 | 1.08 (0.96-1.22) | 0.92 (0.76-1.13) |
| 70–84 beds | 449 (22.0%) | 2040 | 1.26 (1.09-1.44) | 0.94 (0.73-1.21) |
| **Occupants/bedrooms – 0.9\*** |  |  |  |  |
| Mean (SD) |  | 0.9 (0.2) | 0.78 (0.54-1.12) | 0.67 (0.35-1.30) |
| **Beds/staff – 0.85\*** |  |  |  |  |
| Mean (SD) |  | 0.9 (0.2) | 1.32 (1.02-1.70) | 1.36 (0.76-2.45) |
| **Infection/outbreak status** |  |  |  |  |
| **Non-outbreak care homes** |  |  |  |  |
| A Uninfected (other LTCF) | 646 (22.9%) | 2819 | 1.00 (reference) | 1.00 (reference) |
| B Symptomatic not confirmed | 34 (26.0%) | 131 | 4.77 (3.35-6.77) | 4.62 (2.91-7.33) |
| **Outbreak care homes** |  |  |  |  |
| A Uninfected | 636 (13.5%) | 4723 | 2.16 (1.89-2.47) | 2.19 (1.83-2.62) |
| B Symptomatic not confirmed | 161 (37.2%) | 433 | 9.95 (8.21-12.1) | 9.88 (7.01-13.9) |
| C Confirmed asymptomatic | 15 (11.3%) | 133 | 3.68 (2.18-6.20) | 3.84 (2.31-6.40) |
| D Confirmed symptomatic | 202 (42.6%) | 474 | 13.8 (11.5-16.5) | 13.9 (10.8-17.8) |

**Notes:**
Baseline group = uninfected residents in non-outbreak LTCFs
\*HRs of continuous covariates correspond to the effect of an increase by 1 in occupants/bedroom or beds/staff. The effect of a 10 percentage points increase is computed as $HR^{0.1}$ ; for example $60.5^{0.1}=1.5$ is the increase in hazards of infection associated with a 10 percentage points increase in occupancy.

We estimated excess mortality in outbreak and non-outbreak care homes, taking individuals with no evidence of infection (Group A) in non-outbreak care homes as the reference group. Hazards of all-cause mortality were two-fold higher in Group A – no direct evidence of infection in outbreak versus non-outbreak care homes (adjusted HR = 2.2 [1.8; 2.6]). All-cause mortality was strongly associated with confirmed infection, whether asymptomatic (Group C: adjusted HR = 3.8 [2.3; 6.4]) or symptomatic (Group D: adjusted HR = 14 [11; 18]). In confirmed infections, mortality was significantly higher in individuals with a record of symptoms.

### Attributable mortality (individual-level data)

Model-based estimates indicate that 653/1,694 (39%) all-cause deaths were attributable to COVID-19 (Table S7). In care homes with outbreaks only, just 161/1014 (16%) deaths attributable to COVID-19 occurred in people with confirmed infection (Groups C and D).

## Discussion

### Main findings

Across 179 care homes, 22% of residents experienced symptoms while 10% had laboratory-confirmed infections, with a case-fatality rate of 35.7% across the first wave of the pandemic. Residents with no direct evidence of infection in care homes with outbreaks had twice the mortality of the equivalent group in care homes without outbreaks. Only one in six deaths attributable to COVID-19 in outbreak care homes were confirmed due to insufficient testing capacity until late in the pandemic. In addition to the need for active surveillance and increased testing capacity, higher staff-to-resident ratios and reduced occupancy may be important to reduce the spread of infection.

Our estimates are comparable to a large survey of managers of care homes in England.^21^ Both studies are likely to be underestimates due to limited testing, asymptomatic infection^9^ and moderate sensitivity of PCR testing.^22^ Our estimate of 35.7% case-fatality in residents with confirmed infection over a mean 71 days is slightly higher than previous literature,^23–25^ but is based on longer follow-up, a larger number of residents, and our study population had higher overall mortality.

Two-thirds of care homes in our study reported at ≥1 infection or death, in agreement with a study from one region of Scotland which reported that 61% of care homes had experienced an outbreak.^16^ This suggests that most outbreaks were identified through FSHCG’s active surveillance system, and supports our assumption that residents in non-outbreak care homes had not been exposed to infection. This assumption made it possible to estimate mortality attributable to COVID-19.

Our findings of excess deaths in those with no direct evidence of infection may be due to under-ascertainment, direct effects of COVID-19 control measures on delivery of care, and/or indirect effects due to additional disruption caused by the outbreak.^26,27^ Detailed analysis of cause of death and reasons for hospital admission in care home residents will be important to understand how the pandemic has affected the quality of care in care homes. Our analysis provides a method that could be widely applied to estimate excess mortality, provided care homes with outbreaks can be reliably identified.

In common with a Canadian cohort study,^28^ we found strong associations between infections and care home baseline occupancy. We also found staffing levels to be negatively associated with infection rates. These organisational factors, linked to chronic underfunding of the care sector, are likely to hinder the implementation of robust infection control procedures^29^ such as isolating or cohorting infected residents, staff training, and regular environmental deep cleaning. When staff care for fewer residents they also have reduced likelihood of spreading infection between residents. Higher staff-to-resident ratios may also decrease reliance on agency staff working across multiple settings, and indicate better-resourced care homes. These associations may also be confounded by other characteristics which could not be measured in this study, such as access to personal protective equipment, or building structure/layout.

### Strengths and limitations

The unique surveillance system we established in partnership with FSHCG tracked infections across a large number of care homes. To our knowledge, this is the most complete reporting system for COVID-19 infections in care homes published to date. It is possible that care homes that paid less attention to active surveillance to support control will have had higher levels of uncontrolled outbreaks compared to those seen in this study.

Our estimates of mortality attributable to COVID-19 are dependent on our definition of ‘outbreak’ versus ‘non-outbreak’ care homes. We used a sensitive definition (≥1 case/home) due to under-ascertainment caused by the lack of testing. However, it is possible that we incorrectly classified some care homes with only a few cases throughout the pandemic as having experienced outbreaks. Other key limitations relate to the completeness of individual-level data. We lacked information on comorbidity and ethnicity, shown to be important risk factors for adverse outcomes in COVID-19,^4^ but we were able to identify individuals with dementia, and adjust for receipt of nursing care which will partially capture comorbidity. The number of infections was under-reported in the individual-level dataset by comparison with the manager-reported daily infection tallies, and we lacked information on the overall rate of testing in each care home. Finally, our measures of care home occupancy were based on the pre-pandemic period and did not take account of higher vacancy rates during follow-up.

### Conclusions

UK numbers of infected residents and staff were underestimated during the first wave of the pandemic. Our findings support disease control strategies which integrate public health surveillance and rapid testing with investment in care homes to reduce occupancy and increase staffing. Although testing will improve case ascertainment, frequent testing in care home residents may not always be desirable if the risk of infection is low, because the testing procedure (nasopharyngeal swabs) is invasive and may distress vulnerable residents. Since the incubation period and serial interval of COVID-19 is short,^30^ the interval between successive screens required to interrupt transmission may also need to be short. Strengthened surveillance in care homes could be greatly facilitated by the availability of near patient testing platforms, such as lateral flow immunoassays,^31^ provided the predictive value of these tests is adequate.

## Data Availability

All computer syntax used in the analysis is available through an online repository. The data that support the findings of this study are available from the Four Seasons Healthcare Group but restrictions apply to the availability of these data, which were used under a data sharing agreement for the current study and so are not publicly available. Data are however available from the authors upon reasonable request and with permission of Four Seasons Healthcare Group.

https://github.com/peterdutey/ch-covid19

## Ethical approval

This study was approved by the University College London Research Ethics Committee (project reference 13355/002).

## Availability of data and materials

This work uses data provided by residents and collected by the Four Seasons Health Care Group as part of their care and support. These are available from the Four Seasons Health Care Group but restrictions apply to the availability of these data, which were used under license for the current study, and so are not publicly available. Data are however available from the authors upon reasonable request and with permission of the Four Seasons Health Care Group.

## References

1. Public Health England. COVID-19: Number of outbreaks in care homes – management information, https://www.gov.uk/government/statistical-data-sets/covid-19-number-of-outbreaks-in-care-homes-management-information (2020).

2. LaingBuisson. Care of Older People UK Market Report 30th Edition, https://www.laingbuisson.com/ (2019).

3. Sanford AM, Orrell M, Tolson D, et al. An international definition for “nursing home”. Journal of the American Medical Directors Association 2015; 16: 181–184.

4. Zhou F, Yu T, Du R, et al. Clinical course and risk factors for mortality of adult inpatients with COVID-19 in Wuhan, China: a retrospective cohort study. The Lancet 2020; 395: 1054–1062.

5. England Office for National Statistics. Deaths involving COVID-19 in the care sector, England and Wales: deaths occurring up to 12 June 2020 and registered up to 20 June 2020 (provisional), https://www.ons.gov.uk/peoplepopulationandcommunity/birthsdeathsandmarriages/deaths/articles/deathsinvolvingcovid19inthecaresectorenglandandwales/deathsoccurringupto12june2020andregisteredupto20june2020provisional (2020).

6. England Office for National Statistics. Coronavirus (COVID-19) related deaths by occupation, England and Wales: deaths registered between 9 March and 25 May 2020, https://www.ons.gov.uk/peoplepopulationandcommunity/healthandsocialcare/causesofdeath/bulletins/coronaviruscovid19relateddeathsbyoccupationenglandandwales/deathsregisteredbetween9marchand25may2020 (2020).

7. Fisman DN, Bogoch I, Lapointe-Shaw L, et al. Risk factors associated with mortality among residents with coronavirus disease 2019 (covid-19) in long-term care facilities in ontario, canada. JAMA Network Open 2020; 3: e2015957–e2015957.

8. Hollinghurst J, Lyons J, Fry R, et al. The impact of covid-19 on adjusted mortality risk in care homes for older adults in wales, uk: A retrospective population-based cohort study for mortality in 2016–2020. Age and Ageing. Epub ahead of print September 2020. DOI: 10.1093/ageing/afaa207.

9. Salcher-Konrad M, Jhass A, Naci H, et al. COVID-19 related mortality and spread of disease in long-term care: First findings from a living systematic review of emerging evidence. medRxiv. Epub ahead of print 2020. DOI: 10.1101/2020.06.09.20125237.

10. UK Department of Health & Social Care. Press release: Government launches new portal for care homes to arrange coronavirus testing, https://webarchive.nationalarchives.gov.uk/20200616160922/https://www.gov.uk/government/news/government-launches-new-portal-for-care-homes-to-arrange-coronavirus-testing.

11. Smith CM, Williams H, Jhass A, et al. Antibiotic prescribing in UK care homes 2016–2017: retrospective cohort study of linked data. BMC Health Services Research 2020; 20: 555.

12. Abel GA, Barclay ME, Payne RA. Adjusted indices of multiple deprivation to enable comparisons within and between constituent countries of the UK including an illustration using mortality rates. BMJ Open 2016; 6: e012750.

13. England Care Quality Commission. Care directory with filters 2020, https://www.cqc.org.uk/sites/default/files/HSCA_Active_Locations_01_June_2020.xlsx

14. England Office for National Statistics. Coronavirus (COVID-19) Infection Survey pilot: 18 June 2020, https://www.ons.gov.uk/peoplepopulationandcommunity/healthandsocialcare/conditionsanddiseases/bulletins/coronaviruscovid19infectionsurveypilot/latest#covid-19-infection-survey-data.

15. England Office for National Statistics. COVID-19 Infection Survey Datasets, https://www.ons.gov.uk/file?uri=%2fpeoplepopulationandcommunity%2fhealthandsocialcare%2fconditionsanddiseases%2fdatasets%2fcoronaviruscovid19infectionsurveydata%2f2020/covid19infectionsurveydatasets20200618full1.xlsx.

16. Burton JK, Bayne G, Evans C, et al. Evolution and effects of COVID-19 outbreaks in care homes: A population analysis in 189 care homes in one geographical region of the UK. The Lancet Healthy Longevity 2020; 1: e21–e31.

17. Samuelsen SO, Eide GE. Attributable fractions with survival data. Statistics in Medicine 2008; 27: 1447–1467.

18. Aragon TJ. Epitools: Epidemiology tools, https://CRAN.R-project.org/package=epitools (2020).

19. Therneau TM. A package for survival analysis in s, https://CRAN.R-project.org/package=survival (2015).

20. Dutey-Magni P, Shallcross L. COVID-19 infection and attributable mortality in UK long term care facilities: Cohort study using active surveillance and electronic records (March-June 2020). Epub ahead of print 2020. DOI: 10.5281/zenodo.3947780.

21. England Office for National Statistics. Impact of coronavirus in care homes in England (Vivaldi): 26 May to 19 June 2020, https://www.ons.gov.uk/peoplepopulationandcommunity/healthandsocialcare/conditionsanddiseases/articles/impactofcoronavirusincarehomesinenglandvivaldi/26mayto19june2020/previous/v1 (2020).

22. Kucirka LM, Lauer SA, Laeyendecker O, et al. Variation in False-Negative Rate of Reverse Transcriptase Polymerase Chain Reaction–Based SARS-CoV-2 Tests by Time Since Exposure. Annals of Internal Medicine 2020; M20–1495.

23. Blain H, Rolland Y, Tuaillon E, et al. Efficacy of a test-retest strategy in residents and health care personnel of a nursing home facing a covid-19 outbreak. Journal of the American Medical Directors Association 2020; 21: 933–936.

24. McMichael TM, Currie DW, Clark S, et al. Epidemiology of Covid-19 in a Long-Term Care Facility in King County, Washington. New England Journal of Medicine 2020; 382: 2005–2011.

25. Arons MM, Hatfield KM, Reddy SC, et al. Presymptomatic sars-cov-2 infections and transmission in a skilled nursing facility. New England Journal of Medicine 2020; 382: 2081–2090.

26. Lai AG, Pasea L, Banerjee A, et al. Estimating excess mortality in people with cancer and multimorbidity in the covid-19 emergency. medRxiv. Epub ahead of print 2020. DOI: 10.1101/2020.05.27.20083287.

27. Weinberger DM, Chen J, Cohen T, et al. Estimation of Excess Deaths Associated With the COVID-19 Pandemic in the United States, March to May 2020. JAMA Internal Medicine. Epub ahead of print July 2020. DOI: 10.1001/jamainternmed.2020.3391.

28. Brown KA, Jones A, Daneman N, et al. Association between nursing home crowding and covid-19 infection and mortality in ontario, canada. medRxiv. Epub ahead of print 2020. DOI: 10.1101/2020.06.23.20137729.

29. UK Department of Health & Social Care. Admission and Care of Residents in a Care Home during COVID-19. Version 2. Updated 19 June 2020, https://www.gov.uk/government/publications/coronavirus-covid-19-admission-and-care-of-people-in-care-homes (2020).

30. To KK-W, Tsang OT-Y, Leung W-S, et al. Temporal profiles of viral load in posterior oropharyngeal saliva samples and serum antibody responses during infection by SARS-CoV-2: an observational cohort study. The Lancet Infectious Diseases 2020; 20: 565–574.

31. Flower B, Brown JC, Simmons B, et al. Clinical and laboratory evaluation of sars-cov-2 lateral flow assays for use in a national covid-19 seroprevalence survey. Thorax 2020; 75: 1082–1088.

