## Supplementary methods for "COVID-19 infection and attributable mortality in UK care homes: Cohort study using active surveillance and electronic records (March-June 2020)"

Supplementary data quality and methodology report

### 1 Overview

This report provides additional information on the validation of:

- procedures for classifying the SARS-CoV-2 status (symptomatic, tested, confirmed)
- procedures for computing the at-risk resident-days
- overall numbers of symptomatic and confirmed cases

### 2 Classification of SARS-CoV-2 cases from incident reports

The SARS-CoV-2 status is determined by a combination of fields from the Datix incidents management system:

- infection_covid_19_type is a mandatory field one of two possible values:
  - “Confirmed formal clinical diagnosis of COVID-19”
  - “Symptoms, but no definite clinical diagnosis of COVID-19”
- infection_confirmed contains one of 4 possible values (if not missing):
  - “Yes - positive test result: confirmed case”
  - “Test result not yet received”
  - “No - negative test result”
  - “Not tested”
- infection_result is a free-text field containing, for the most part, the words ‘positive’ or ‘negative’

Incident reports are dynamic: they are designed to be updated as more information becomes available. If all fields are not accurately kept updated, some values may become inconsistent. Inconsistencies are observed in incident reports as illustrated in the tabulations below. Due to this, a reclassification algorithm is used which prioritises any information on a positive test result.

|  | | infection_covid_19_type | | | Total |
| --- | --- | --- | --- | --- | --- |
|  |  | Confirmed formal clinical diagnosis of COVID-19 | Symptoms, but no definite clinical diagnosis of COVID-19 | *Missing* |  |
| infection_confirmed |  |  |  |  |  |
| No - negative test result |  | 10  (4.3%) | 225  (95.7%) | 0 (0.0%) | 235 (100.0%) |
| Not tested |  | 4  (1.1%) | 366  (98.9%) | 0 (0.0%) | 370 (100.0%) |
| Test result not yet received |  | 2  (1.6%) | 121  (98.4%) | 0 (0.0%) | 123 (100.0%) |
| Yes - positive test result: confirmed case |  | 69  (92.0%) | 60  (8.0%) | 0 (0.0%) | 751 (100.0%) |
| *Missing* |  | 0  (0.0%) | 13  (100.0%) | 0 (0.0%) | 13 (100.0%) |
| Total |  | 707  (47.4%) | 785  (52.6%) | 0 (0.0%) | 1,492 (100.0%) |

#### 2.1 Algorithm

The following algorithm is used to classify symptomatic, tested, and test-confirmed cases from the Datix reports of COVID-19 infections.

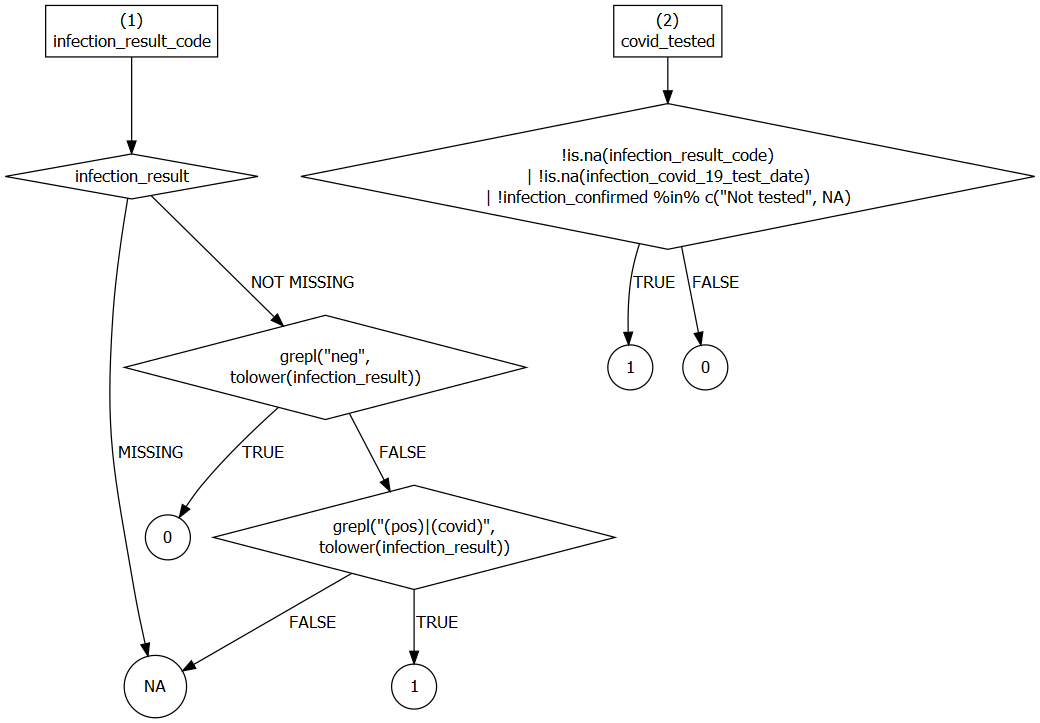

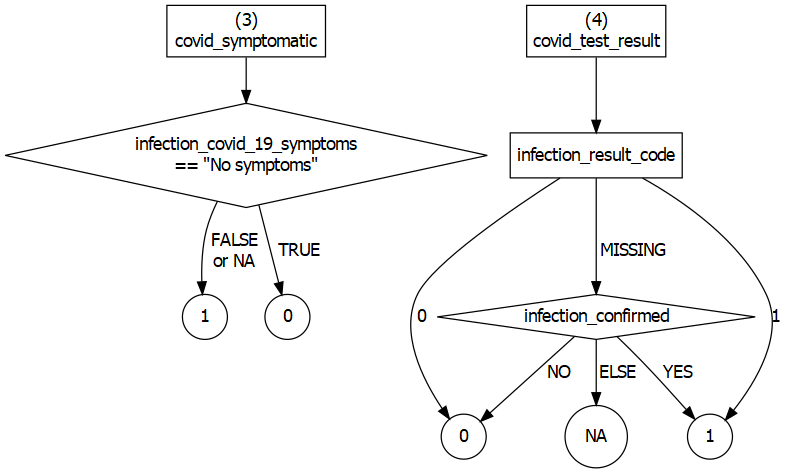

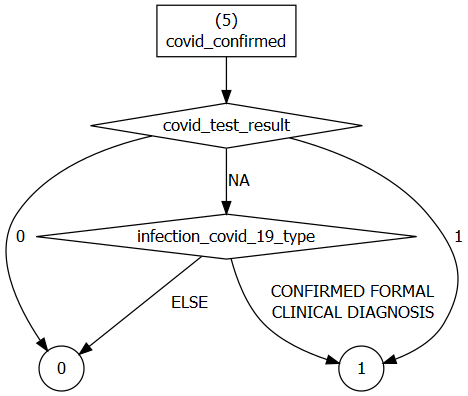

#### 2.2 Validation

Data are tabulated below to evaluate the internal consistency of the rules set out in the previous section.

The infection_result field provides information on test results which broadly agrees yet sometimes conflicts with the infection_confirmed classification of incidents. It is potentially more up to date although it is very sparsely populated.

|  | covid_confirmed | | | |
| --- | --- | --- | --- | --- |
| infection_covid_19_type | 0 | 1 | *Missing* | Total |
| Confirmed formal clinical diagnosis of COVID-19 | 11  (1.6%) | 696  (98.4%) | 0  (0.0%) | 707  (100.0%) |
| Symptoms, but no definite clinical diagnosis of COVID-19 | 720  (91.7%) | 65  (8.3%) | 0  (0.0%) | 785  (100.0%) |
| *Missing* | 0  (0.0%) | 0  (0.0%) | 0  (0.0%) | 0  (0.0%) |
| Total | 731  (49.0%) | 761  (51.0%) | 0  (0.0%) | 1,492 (100.0%) |

|  | covid_test_result | | | |
| --- | --- | --- | --- | --- |
| infection_covid_19_type | 0 | 1 | *Missing* | Total |
| Confirmed formal clinical diagnosis of COVID-19 | 11  (1.6%) | 693  (98.0%) | 3  (0.4%) | 707  (100.0%) |
| Symptoms, but no definite clinical diagnosis of COVID-19 | 226  (28.8%) | 65  (8.3%) | 494  (62.9%) | 785  (100.0%) |
| *Missing* | 0  (0.0%) | 0  (0.0%) | 0  (0.0%) | 0  (0.0%) |
| Total | 237  (15.9%) | 758  (50.8%) | 497  (33.3%) | 1,492  (100.0%) |

|  | covid_test_result | | | |
| --- | --- | --- | --- | --- |
| infection_confirmed | 0 | 1 | *Missing* | Total |
| No - negative test result | 235  (100.0%) | 0  (0.0%) | 0  (0.0%) | 235  (100.0%) |
| Not tested | 0  (0.0%) | 6  (1.6%) | 364  (98.4%) | 370  (100.0%) |
| Test result not yet received | 1  (0.8%) | 2  (1.6%) | 120  (97.6%) | 123  (100.0%) |
| Yes - positive test result: confirmed case | 1  (0.1%) | 750  (99.9%) | 0  (0.0%) | 751  (100.0%) |
| *Missing* | 0  (0.0%) | 0  (0.0%) | 13  (100.0%) | 13  (100.0%) |
| Total | 237  (15.9%) | 758  (50.8%) | 497  (33.3%) | 1,492  (100.0%) |

### 3 Data quality

#### 3.1 Data linkage

Secondary use individual-level data for private residents receiving general and elderly care were extracted and pseudonymised by HW. They consisted of two datasets:

- **a residents dataset** containing one record per resident per care home, alongside residents’ gender, date of birth, most recent dates of admission and discharge, date of first admission, type of stay (residential/nursing) and care (general, dementia, elderly). This dataset does not include any records for local authority ‘block contract’ beds.
- **an incidents dataset** containing 1,880 reports filed by care home staff. Mandated report field were: resident forename, surname, care home identifier, incident date/time, and date/time of reporting. In addition, reports could record: date of birth, gender, information on Covid-19 symptoms, tests and test results, resident current location (home/hospital), and death. Incidents were classified to indicate whether the resident was symptomatic, and whether an infection was confirmed on the basis of multiple variables (supplementary material 1).

HW created a resident index file bridging the two datasets using: the first three letters of the resident’s forename; the resident’s full surname; and the care home identifier. Clerical review of identifiers generated in this way established that no single identifier was allocated to distinct individuals in the residents’ dataset. Just 65 (3.5%) incident records did not contain enough information to generate an identifier and were excluded. Identifiers of a further 323 records (17.2%) could not be successfully linked to the residents dataset and were excluded, after clerical review established they could not be matched with a resident. A proportion of these records may have been filed in error, while others may re-late to occupants of local authority beds who are not accounted for in the residents’ dataset.

#### 3.2 Exclusions

A total of 1,880 incident reports were obtained. Resident pseudoidentifiers were absent in 65 (3.5%) of Datix incident reports. The pseudoidentifiers of a further 323 reports (17.2%) were could not be successfully linked to the residents database. All were excluded, leaving a total of 1492 reports dated between Monday 2 March 2020 and Sunday 14 June 2020 relating to 1,206 unique residents. These reports belongued to 126 care homes totalling 6,964 beds. For the period of study, the Four Seasons Health Care Group (FSHCG) comprises 179 care homes with a total of 9,568 beds.

#### 3.3 Occupancy

Care home resident-days are available from two sources:

- reports to FSHCG by every home on their occupancy every Sunday, which include residents occupying contract beds
- individual-level resident records of first and last admission and discharge dates, to he exclusion of occupants of contract beds. These records are used to compute occupancy on a daily basis.

In total, 855 out of 9,568 beds were contracted by the local authority, with 6 homes having more than half of their beds contracted.

Plots below compare both measures for every Sunday (Figure 1) and in total across the period (Figure 2) at the level of care homes, drawing attention to care homes with a large proportion of contract beds. The Pearson’s correlation between the weekly measures was $r=$ 0.77 overall, rising to $r=$ 0.97 once excluding care homes with $\geq$ 10% contract beds. These findings confirm that occupancy could be approximated reliably using just residents’ records of first and last admission and discharge dates.

This enabled us to estimate exposure (resident-days) excluding local authority contract beds, since no Datix incident reports were returned for occupants of such beds.

In addition, this validation enabled us to approximate the total number of unique residents (including contract beds) during the period of observation for aggregate count data (2020-03-24 – 2020-06-14). The ratio of occupants recorded by FSHCG to the number approximated by us was 1.140. The number of unique residents in non-contract beds was 8,713 during the study period. By assuming that the resident turnover in local authority beds was the same as the rate in other beds, we infer the number of unique residents inclusive of contract beds to be 9,931.

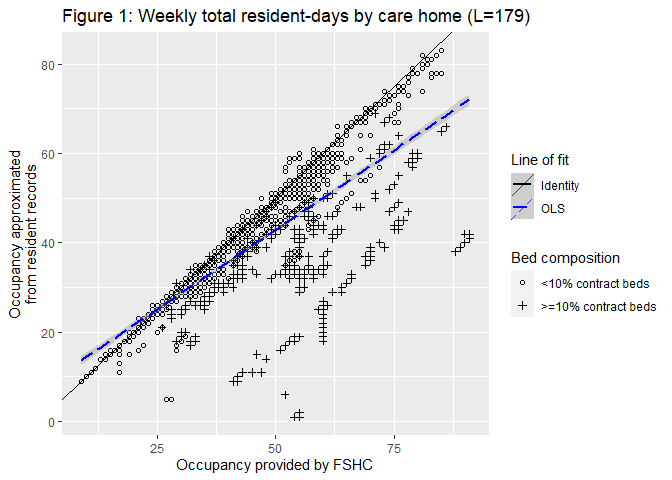

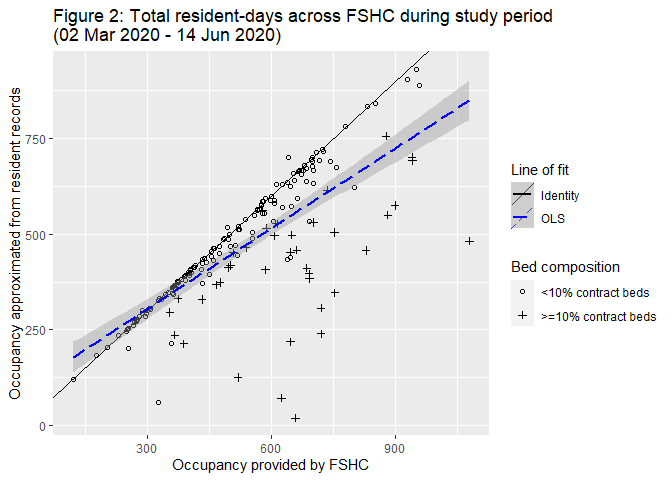

We nevertheless identified 11 care homes in which the total resident-days approximated differed by more than 10 percent despite not belonging (Figure 2).

#### 3.4 Comparison of manager-reported tally counts with Datix-derived counts

During the period, manager counted 951 confirmed infection while Datix recorded 605 such infections. The 57% excess could be due to a range of explanations:

1. contract beds occupants, who may represent at least 12.3% of residents (assuming that their turnover is identical to that of occupants of other beds, in reality turnover is likely to be higher, and so is this proportion), may have a higher infection rate than other residents
2. underreporting on Datix
3. linkage error
4. double-counting of cases in manager counts

It is not possible to rely on explanations (1-3) exclusively: the number of confirmed infections on Datix was greater than manager counts in 19 care homes. This suggest double counting (4) is a possibility.

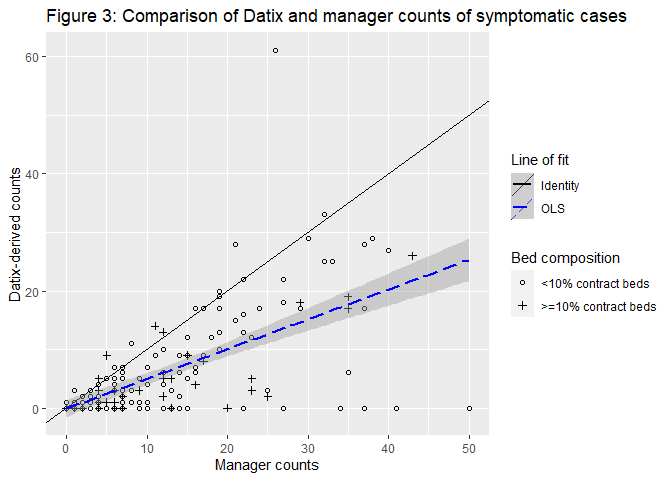

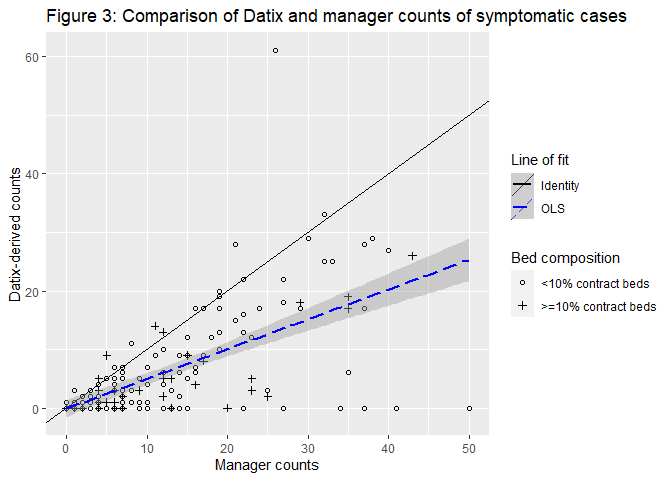

### 4 Estimating the cumulative incidence of cases and resident-days at risk

Manager-reported outbreak surveillance counts were used together with occupancy estimated from weekly occupied bed censuses in order to estimate the incidence of cases across time using a Kaplan-Meier life table product limit estimator.

The life table records for every day $t=\{0,\ldots,T\}$:

- $n_{t}$, the number of residents present in the home on day $t$
- $I_{t}$, the number of residents infected up to and including day $t-1$ who are still present in the home
- $d_{t}$, the number of residents with a new infection on day $t$ (includes residents admitted on day $t$ who got infected outside the home, for instance in hospital)
- $S_{t}$, the number of exposed-to-risk residents, that is, the number of residents who have not yet had an infection and are susceptible to an infection.

Note that on any day, $n_{t}=S_{t}+I_{t}$.

Neither $S_{t}$ or $I_{t}$ were recorded as such by care homes. Instead, these numbers were approximated in a multiple decrement life table. For this, we assumed that:

- no residents were infected before 24 March 2020 ($I_{t=0}=0$) when tallies began
- residents previously infected (counted in $I_{t}$) are discharged at the same rate as exposed-to-risk residents (counted in $S_{t}$). This assumption was verified in the individual-level records.

Admitted cases were accounted for in the daily new case count. Based on this

Based on these assumptions, we estimated $I_{t}$ and $S_{t}$ as

$$\hat{I_{t}}=min(n_{t}/n_{t-1},1)(\hat{I}_{t-1}+d_{t-1}); I_{t=0}=0$$

$$\hat{S_{t}}=n_{t}-\hat{I_{t}}$$

where $min(n_{t}/n_{t-1},1)$ serve as an (underestimated) approximation of the discharge rate.

The cumulative exposure time (used for computing cumulative incidence) was calculated as $\sum_{t=0}^{T} \hat{S}_{t}$.

### 5 Total homes, beds, contract beds, and residents within FSHCG and nationwide

|  |  | **FSHCG** |  |  | **Nationwide** |
| --- | --- | --- | --- | --- | --- |
|  | **Total homes** | **Total beds** |  | **Total homes** | **Total beds** |
| England | 112 | 6,085 |  | 9,400 | 45,000 |
| Northern Ireland | 43 | 2,074 |  | 483 | 16,095 |
| Scotland | 24 | 1,409 |  | 1,142 | 40,926 |

Data sources: (England Care Quality Commission 2020; Northern Ireland Health and Social Care Board 2020; NHS National Services Scotland Information Services Division 2020)

### 6 Sensitivity analysis

This section reports an alternative set of estimates based on a narrower definition of ‘outbreak’ versus ‘non-outbreak’ homes than was used in the paper: here ‘outbreak homes’ are defined as homes recording ≥2 confirmed cases of infection in the manager-reported aggregate data.

Two tables are sensitive to this definition:

- Risk factors for all-cause mortality (Table 6)
- Model-based estimates of attributable death (Table S7).

We found that the narrow definition (≥2 confirmed cases), only 69/179 (39%) homes were classified as outbreak homes by the end of the study period, compared to 121/179 (68%) homes with more sensitive definition used in the paper.

Using the narrow definition, just 438/1,694 all-cause deaths (26%) in private residents were attributable to COVID-19, compared to 653/1,694 (39%) with the more sensitive definition used in the paper.

Table 6. Sensitivity to ‘outbreak definition’: Risk factors for all-cause mortality in private residents of care homes with and without COVID-19 outbreaks: hazard ratios (HR) from a Cox proportional hazards model (n=8,713, 2 Mar 2020-14 Jun 2020)

|  | Deaths | N | HR (univariate) | HR (multivariate) |
| --- | --- | --- | --- | --- |
| **Gender** |  |  |  |  |
| Female | 996 (17·6%) | 5667 | 1.00 (reference) | 1.00 (reference) |
| Male | 698 (22·9%) | 3046 | 1.40 (1.27-1.54) | 1.46 (1.32-1.62) |
| **Age** |  |  |  |  |
| <75 years | 201 (14·1%) | 1424 | 1.00 (reference) | 1.00 (reference) |
| 75–84 years | 520 (18·2%) | 2865 | 1.30 (1.11-1.53) | 1.35 (1.13-1.61) |
| 85–94 years | 761 (21·0%) | 3629 | 1.50 (1.28-1.75) | 1.71 (1.46-2.01) |
| 95+ years | 212 (26·7%) | 795 | 1.93 (1.59-2.34) | 2.28 (1.83-2.84) |
| **Bed type** |  |  |  |  |
| Residential | 420 (15·4%) | 2734 | 1.00 (reference) | 1.00 (reference) |
| Nursing | 1274 (21·3%) | 5979 | 1.38 (1.24-1.54) | 1.33 (1.11-1.60) |
| **Care type** |  |  |  |  |
| General/elderly | 1015 (19·2%) | 5294 | 1.00 (reference) | 1.00 (reference) |
| Dementia | 679 (19·9%) | 3419 | 1.02 (0.92-1.12) | 1.00 (0.86-1.16) |
| **Index of Multiple Deprivation** |  |  |  |  |
| 1 - least deprived | 469 (50.1%) | 937 | 0.99 (0.84-1.17) | 1.05 (0.79-1.39) |
| 2 | 190 (12.6%) | 1508 | 0.85 (0.73-0.99) | 0.83 (0.62-1.10) |
| 3 (reference category) | 266 (11.5%) | 2320 | 1.00 (reference) | 1.00 (reference) |
| 4 | 321 (19.1%) | 1680 | 0.92 (0.80-1.06) | 0.85 (0.67-1.10) |
| 5 - most deprived | 448 (19.8%) | 2268 | 0.98 (0.86-1.12) | 0.90 (0.71-1.14) |
| **Total beds** |  |  |  |  |
| 20–34 beds | 373 (17.5%) | 2129 | 1.00 (reference) | 1.00 (reference) |
| 45–59 beds | 872 (19.2%) | 4544 | 1.08 (0.96-1.22) | 0.95 (0.78-1.17) |
| 70–84 beds | 449 (22.0%) | 2040 | 1.26 (1.09-1.44) | 1.05 (0.82-1.35) |
| **Occupants/bedrooms – 0.9*** |  |  |  |  |
| Mean (SD) |  | 0.9 (0.2) | 0.78 (0.54-1.12) | 0.72 (0.36-1.40) |
| **Beds/staff – 0.85*** |  |  |  |  |
| Mean (SD) |  | 0.9 (0.2) | 1.32 (1.02-1.70) | 1.54 (0.89-2.64) |
| **Infection/outbreak status** |  |  |  |  |
| **Non-outbreak care homes** | | | | |
| A Uninfected | 994 (19.5%) | 5092 | 1.00 (reference) | 1.00 (reference) |
| B Symptomatic not confirmed | 92 (39.3%) | 234 | 5.78 (4.65-7.19) | 5.69 (3.93-8.24) |
| **Outbreak care homes** | | | | |
| A Uninfected | 288 (11.8%) | 2450 | 1.86 (1.60-2.15) | 1.86 (1.45-2.39) |
| B Symptomatic not confirmed | 103 (31.2%) | 330 | 6.82 (5.50-8.45) | 6.48 (4.37-9.62) |
| C Confirmed asymptomatic | 15 (11.3%) | 133 | 2.74 (1.64-4.60) | 2.79 (1.67-4.65) |
| D Confirmed symptomatic | 202 (42.6%) | 474 | 10.4 (8.82-12.2) | 10.3 (7.93-13.4) |

*Notes:*
Baseline group = uninfected residents in non-outbreak care homes

*HRs of continuous covariates correspond to the effect of an increase by 1 in occupants/bedroom or beds/staff. The effect of a 10 percentage points increase is computed as HR^0.1^; for example 60.5^0.1^=1.5 is the increase in hazards of infection associated with a 10 percentage points increase in occupancy.

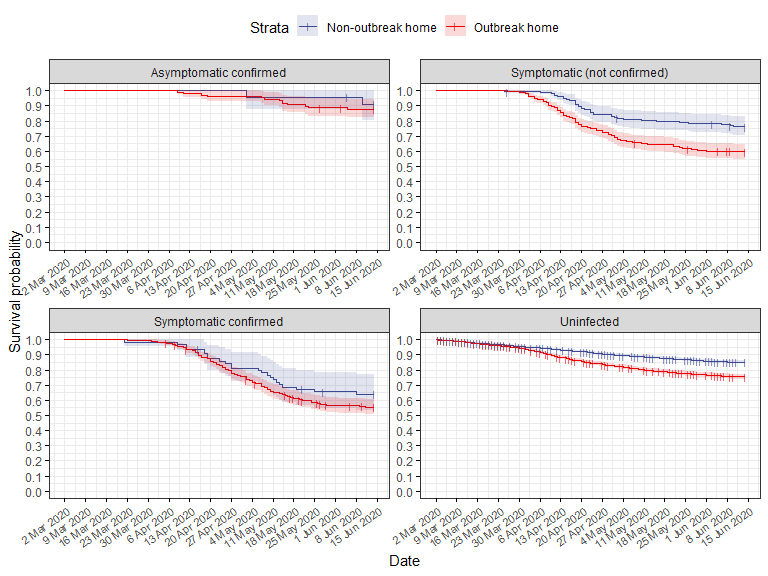

**Figure S3** Sensitivity to ‘outbreak definition’: Kaplan-Meier estimates of resident (n=8,713) survival by SARS-COV-2 case type

**Table S7** Sensitivity to ‘outbreak definition’: Model-based estimates of attributable death in private residents of care homes with and without COVID-19 outbreaks (n=8,713, 2 Mar 2020-14 Jun 2020)

| **Infection/outbreak status** | | **Adjusted HR** | **Resident-days** | **Deaths  attributable to COVID-19** | **All-cause deaths** | % |
| --- | --- | --- | --- | --- | --- | --- |
|  | **Non-outbreak care homes** | | | | | |
| A Uninfected | | 1.00 [1.00; 1.00] | 584,809.57 | 0 | 994 | 0.00^#^ |
| B Symptomatic not confirmed | | 5.69 [3.93; 8.24] | 10,265.40 | 67 | 92 | 15.3^#^ |
|  | **Outbreak care homes** | | | | | |
| A Uninfected | | 1.86 [1.45; 2.39] | 119,841.75 | 132 | 288 | 30.1^#^ |
| B Symptomatic not confirmed | | 6.48 [4.37; 9.62] | 12,341.08 | 77 | 103 | 17.6^#^ |
| C Confirmed asymptomatic | | 2.79 [1.67; 4.65] | 5,300.03 | 10 | 15 | 2.3^#^ |
| D Confirmed symptomatic | | 10.3 [7.93; 13.4] | 14,933.59 | 153 | 202 | 34.9^#^ |
| TOTAL | |  | **747,491** | **438** | **1,694** | **25.9*** |

*Note:* #: column percent; *: row percent

### References

England Care Quality Commission. 2020. “Care Directory with Filters.” <https://www.cqc.org.uk/sites/default/files/HSCA_Active_Locations_01_June_2020.xlsx>.

NHS National Services Scotland Information Services Division. 2020. “Care Home Census for Adults in Scotland Figures for 2007-2017 (as at 31 March).” <https://www.isdscotland.org/Health-Topics/Health-and-Social-Community-Care/Publications/2018-09-11/2018-09-11-CHCensus-Report.pdf?13142031432>.

Northern Ireland Health and Social Care Board. 2020. “Coronavirus (COVID-19) Care Homes - Surge Plan and Frequently Asked Questions.” <http://www.hscboard.hscni.net/coronavirus/covid-19-care-homes/>.
