## Supplementary tables and figures for "COVID-19 infection and attributable mortality in UK care homes: Cohort study using active surveillance and electronic records (March-June 2020)"

**Table S1a** Cumulative incidence and rate of SARS-CoV-2 infections in residents according to FSHCG aggregate data (2 Mar 2020-14 Jun 2020)

| **Residents** | **All care homes** | | |  | **Outbreak care homes only** | | |
| --- | --- | --- | --- | --- | --- | --- | --- |
|  | **Symptomatic** | **Confirmed** | **COVID-19**  **related deaths** |  | **Symptomatic** | **Confirmed** | **COVID-19**  **related deaths** |
| Cases | 2,075 | 951 | 526 |  | 1,807 | 951 | 526 |
| $N$ exposed | 9,339 | 9,339 | 9,339 |  | 7,102 | 7,102 | 7,102 |
| Total exposure (days) | 563,901 | 623,161 | 659,843 |  | 383,536 | 430,133.8 | 466,813 |
| Cumulative incidence (%) | 22.2  [21.4; 23.1] | 10.2  [9.6; 10.8] | 5.6  [5.2; 6.1] |  | 25.4  [24.4; 26.5] | 13.4  [12.6; 14.2] | 7.4  [6.8; 8.0] |
| Incidence rate (per 100,000  person-days) | 368.0  [352.3; 384.2] | 152.6  [143.1; 162.6] | 79.7  [73.0; 86.8] |  | 471.1  [449.7; 493.4] | 221.1  [207.3; 235.6] | 112.7  [103.3; 122.7] |

**Table S1b** Incidence proportions and rates of SARS-CoV-2 infections amongst residents according to Datix incident reports (2 Mar 2020–14 Jun 2020)

| **Residents** | **Symptomatic** | **Confirmed** | All-cause death |
| --- | --- | --- | --- |
| Cases | 1,045 | 617 | 1,694 |
| $N$ exposed | 8,713 | 8,713 | 8,713 |
| Total exposure (days) | 708,787 | 726,009 | 746,090 |
| Cumulative incidence (%) | 12.0 [11.3; 12.7] | 7.1 [6.6; 7.6] | 19.4 [18.6; 20.3] |
| Incidence rate (per 100,000) | 147.4 [138.6; 156.7] | 85.0 [78.4; 92.0] | 227.1 [216.4; 238.1 ] |

Datix incidents reports recorded that 928 private residents were tested at least once. Test results were known for 828 of them (89.2%), including 617 residents with a positive test result. This data should be treated as representative, as tests coming back negative were not commonly recorded on Datix. Incidence proportions and rates reported in Table S1B above are lower than corresponding estimates based on outbreak counts. This is thought to be caused by a combination of under-ascertainment as well as linkage error.

**Table S2** Cumulative incidence and rate of SARS-CoV-2 infections among staff according to FSHCG aggregate data (2 Mar 2020-14 Jun 2020)

| **Staff** | **Symptomatic** | Confirmed |
| --- | --- | --- |
| Cases | 1,892 | 585 |
| $N$ exposed | 11,604 | 11,604 |
| Total exposure (days) | 856,323 | 939,312 |
| Cumulative incidence (%) | 16.3 [15.6; 17.0] | 5.0 [4.7; 5.5] |
| Incidence rate (per 100,000 person-days) | 220.9 [211.1; 231.1] | 62.3 [57.3; 67.5] |

**Table S4** All-cause case-fatality rates by age and sex among private residents (n=8,713; 2 Mar 2020-14 Jun 2020)

| **Age** | **Sex** | **N** | **Confirmed infections** | **Total deaths** | **Deaths in confirmed**  **infections** | Case-fatality  rate (%) |
| --- | --- | --- | --- | --- | --- | --- |
| <75 years | Female | 712 | 48 | 87 | 9 | 18.8 [8.9; 32.6] |
| 75–84 years | Female | 1,687 | 114 | 254 | 31 | 27.2 [19.3; 36.3] |
| 85–94 years | Female | 2,617 | 173 | 484 | 58 | 33.5 [26.5; 41.1] |
| 95+ years | Female | 651 | 42 | 171 | 18 | 42.9 [27.7; 59.0] |
| <75 years | Male | 712 | 37 | 114 | 12 | 32.4 [18.0; 49.8] |
| 75–84 years | Male | 1,178 | 96 | 266 | 41 | 42.7 [32.7; 53.2] |
| 85–94 years | Male | 1,012 | 86 | 277 | 44 | 51.2 [40.1; 62.1] |
| 95+ years | Male | 144 | 11 | 41 | 4 | 36.4 [10.9; 69.2] |
| All | All | 8,713 | 607 | 1,694 | 217 | 35.7 [31.9; 39.7] |

**Table S7** Model-based estimates of attributable death in private residents of care homes with and without COVID-19 outbreaks (n=8,713, 2 Mar 2020-14 Jun 2020)

| **Infection/outbreak status** | | **Adjusted HR** | **Resident-days** | **Deaths  attributable to COVID-19** | **All-cause deaths** | % |
| --- | --- | --- | --- | --- | --- | --- |
|  | **Non-outbreak care homes** | | | | | |
| A Uninfected | | 1.00 [1.00; 1.00] | 436,413 | 0 | 646 | 0.0^#^ |
| B Symptomatic not confirmed | | 4.62 [2.91; 7.33] | 6,382 | 25 | 34 | 3.8^#^ |
|  | **Outbreak care homes** | | | | | |
| A Uninfected | | 2.19 [1.83; 2.62] | 268,178 | 343 | 636 | 52.5^#^ |
| B Symptomatic not confirmed | | 9.88 [7.01; 13.9] | 16,235 | 124 | 161 | 19.0^#^ |
| C Confirmed asymptomatic | | 3.84 [2.31; 6.40] | 5,313 | 11 | 15 | 1.7^#^ |
| D Confirmed symptomatic | | 13.9 [10.8; 17.8] | 15,028 | 150 | 202 | 23.0^#^ |
| TOTAL | |  | 747,549 | 653 | 1,694 | 38.5* |

*Note:* #: column percent; *: row percent

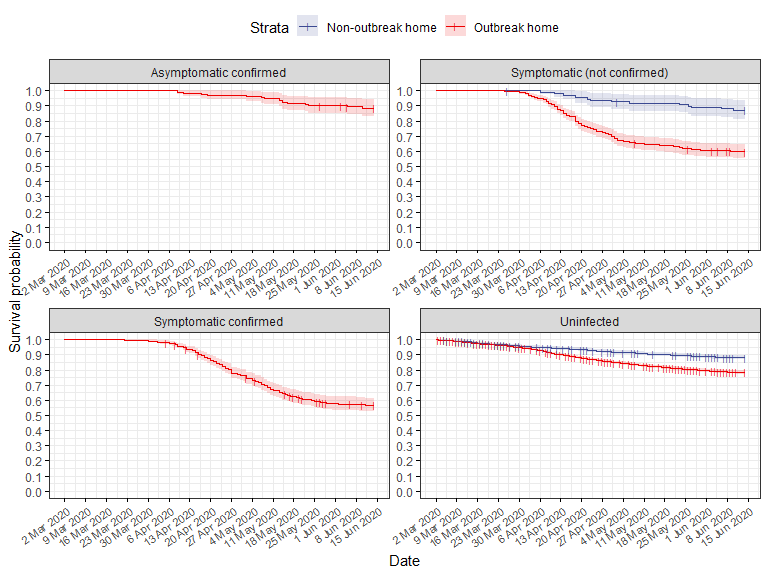

**Figure S3** Kaplan-Meier estimates of resident (n=8,713) survival by SARS-COV-2 case type
